# Health-seeking behavior and barriers to tuberculosis diagnosis and treatment in rural Gurugram, India: A mixed-methods study

**DOI:** 10.64898/2025.12.01.25341399

**Authors:** Vineet Kumar Pathak, Madhulekha Bhattacharya, Namrta Jha, Rohit Sokhey, Apila, Saparya Tripathi

## Abstract

**Background:** The public health problem of tuberculosis (TB) in India worsens because of extended time needed for diagnosis and treatment initiation which affects rural areas most. The National TB Elimination Program (NTEP) requires information about patient healthcare search patterns and their barriers to access in order to achieve its goals.

**Objective:** The research studied rural Gurugram Haryana medical care usage patterns and TB diagnosis and treatment barriers and enablers through patient and healthcare provider feedback collection.

The research employed a mixed-methods cross-sectional design to study 201 TB patients who enrolled on the Nikshay Portal between November 2021 and October 2022 at CHC Farukhnagar. The research team conducted data analysis of semi-structured interview data through univariate and multivariate regression methods using STATA SE software. The research team conducted five focused group discussions (FGDs) across villages to analyze qualitative data through NVivo software for thematic analysis.

**Results:** The participants’ average age reached 35 years while their standard deviation reached 15.3 years. The study participants included 56.7% people who were 35 years old or younger and 53.2% of the participants were male. The study found that 58.2% of participants showed good health-seeking behavior which depended on their knowledge that TB is infectious (OR 2.78, 95% CI 1.39–5.54, p=0.003) and their decision to visit health facilities after home remedies failed (OR 13.07, 95% CI 6.63–25.7, p<0.001). The study found that patients faced three main obstacles to receive proper diagnosis which included delayed medical evaluation and financial constraints and insufficient knowledge about smoking-related TB risks. The qualitative research results showed that patients faced three main barriers because of social discrimination and insufficient assistance and their preference for seeking medical care at private facilities.

**Conclusion:** The improvement of health-seeking behavior depends on TB transmission education and public healthcare facility accessibility and reduced patient expenses and better access to public healthcare facilities. The research data enables NTEP to create successful plans which will help eliminate TB from rural Indian communities by 2025.

## Introduction

The infectious disease Tuberculosis (TB) which Mycobacterium tuberculosis causes has affected human populations for more than 4000 years. The Global TB Report 2021 [1] shows that TB continues to be a major health threat in India because it affects 188 people out of 100,000 population annually in 2020. The National Strategic Plan for TB Elimination [2] supports the National TB Elimination Program (NTEP) which developed from the Revised National TB Control Program (RNTCP) to achieve TB elimination by 2025. The National TB Elimination Program (NTEP) provides free diagnostic and treatment services yet passive case detection continues because patients select healthcare facilities based on their health-seeking behavior (HSB).

The rural Indian population develops their health-seeking behavior (HSB) through their social background and their understanding of diseases and their experience with discrimination and their ability to access medical facilities. Research shows that between 50% and 80% of TB patients visit private medical facilities before they get inadequate treatment at high costs in these facilities [3–4]. The public health services of NTEP do not stop patients from choosing private care because they lack awareness and trust in the system. The post-COVID-19 period has made these problems worse because patients are leaving treatment and drug-resistant TB cases are increasing which proves the need to evaluate how people seek medical care and what blocks their access to treatment. The rural section of Gurugram in Haryana shows these problems because it contains farming communities together with new urban construction. The research used a mixed-methods design to achieve two main objectives by analyzing HSB data quantitatively and FGD results qualitatively to gain complete understanding of TB care systems.

## Materials and Methods

The research used a cross-sectional mixed-methods design which combined quantitative survey data with qualitative FGDs to study HSB and TB management challenges. The research took place in rural Gurugram Haryana at the Community Health Centre (CHC) Farukhnagar field practice site of SGT Medical College. The researchers chose this rural area because it demonstrates the characteristics of the underserved Indian population which NTEP aims to serve. The study included 201 TB patients who received treatment at SGT Medical College from November 2021 through October 2022 and lived within 15 kilometers of the hospital. The research included TB patients who completed their minimum six months of treatment and resided within the study area. The research excluded participants who chose not to participate or failed to provide data or could not be reached despite three attempts at contact. The researchers performed complete enumeration to achieve representation without any sampling errors. The researchers conducted five FGDs with patients from different villages based on their location and TB case distribution.

### Data Collection

#### Quantitative Component

The research team created an interviewer-administered questionnaire which they tested with 20 participants (10 males and 10 females) before making final adjustments. The survey contained three sections which obtained information about (1) personal background details and (2) barriers patients encountered during their diagnosis and treatment process and (3) their medical treatment costs. The researchers made contact with participants through phone calls and video meetings before they gave their verbal consent. The researchers obtained caregiver permission for all participants under 18 years old(assent/ parental consent). The researchers collected data from January 2023 through October 2023 during a 10-month period.

#### Qualitative Component

The researchers conducted five FGDs with 8 to 12 participants from each village who used Hindi and Haryanvi languages for recording and English translation of the transcripts. The FGD guide asked participants to share their experiences before diagnosis and their treatment process and their financial struggles and their experiences with stigma and their interactions with healthcare providers. The trained moderators created a discussion area which enabled participants to share their thoughts freely without any limitations.

The researchers used a scoring system which ranged from 0 to 8 to evaluate HSB through three components: healthcare facility usage and provider selection and diagnostic procedure length. The research analyzed eight independent factors which included age and sex and education level and occupation type and substance abuse and TB knowledge and healthcare service usage patterns. The researchers used odds ratios (OR) to evaluate both barriers and enablers and to determine qualitative themes.

### Data Analysis

#### Quantitative

Data were entered into MS Excel, cleaned, and analyzed using STATA SE. Descriptive statistics (means, SD, frequencies) characterized the sample. Univariate logistic regression identified associations with HSB, followed by multivariate regression for significant variables (p<0.05). Tests of significance (chi-square, t-tests) assessed differences.

#### Qualitative

Transcripts were imported into NVivo for thematic analysis. Codes were developed inductively, grouped into themes (e.g., pre-diagnosis, treatment challenges), and triangulated with quantitative findings.

### Ethical Considerations

Ethical clearance was obtained from the SGT University Institutional Ethics Committee. Verbal consent was secured, confidentiality maintained, and participants could withdraw anytime. Sick patients or defaulters identified were referred to CHC Farukhnagar.

## Results

### Quantitative Findings

The mean (SD) age of study participants was 35 (15.3) years with the range from 4 to 85 years. Among then nearly 56.7% (n=114) were aged ≤35 years and 43.3% (n=87) >35 years. Males comprised 53.2% (n=107), and females 46.8% (n=94). Occupationally, 42.3% (n=85) were not working, 41.3% (n=83) were employed, and 16.4% (n=33) were students. In educational status around 11.9% (n=24) were illiterate, 11.4% (n=23) had up to 5th-grade education, 27.8% (n=76) 6–10th grade, 23.4% (n=47) 11–12th grade, and 15.4% (n=31) were graduates or higher. Majority of them, around 61.2%, (n=123) lived in nuclear families and 38.8% (n=78) in joint families. Smokeless tobacco use was reported by 11.4% (n=23) currently and 27.9% (n=56) in the past. Current smoked tobacco use was 11.9% (n=24), with 31.9% (n=64) reporting past use. Alcohol use was less common, with 8.5% (n=17) current and 23.9% (n=48) past users “Table 1”.

**Table 1:** Distribution of Study Participants by Age, Socio-Demographic Characteristics, and Substance Use (n=201)

| Variable | Category | Frequency (%) |
| --- | --- | --- |
| Age | ≤35 years | 114 (56.7) |
|  | >35 years | 87 (43.3) |
| Sex | Male | 107 (53.2) |
|  | Female | 94 (46.8) |
| Occupation | Not working | 85 (42.3) |
|  | Student | 33 (16.4) |
|  | Employed | 83 (41.3) |
| Education | Illiterate | 24 (11.9) |
|  | Up to 5th | 23 (11.4) |
|  | 6–10th | 76 (27.8) |
|  | 11–12th | 47 (23.4) |
|  | Graduate+ | 31 (15.4) |
| Family Type | Nuclear | 123 (61.2) |
|  | Joint | 78 (38.8) |
| Current Smokeless Tobacco | No | 178 (88.6) |
|  | Yes | 23 (11.4) |
| Past Smokeless Tobacco | No | 145 (72.1) |
|  | Yes | 56 (27.9) |
| Current Smoked Tobacco | No | 177 (88.1) |
|  | Yes | 24 (11.9) |
| Past Smoked Tobacco | No | 137 (68.2) |
|  | Yes | 64 (31.9) |
| Current Alcohol | No | 184 (91.5) |
|  | Yes | 17 (8.5) |
| Past Alcohol | No | 153 (76.1) |
|  | Yes | 48 (23.9) |

The results showed that participants had different levels of TB awareness as 23.9% (n=48) understood smoking leads to TB while 75.1% (n=151) identified symptoms and 78.1% (n=157) recognized TB transmission and 69.4% (n=140) confirmed TB can be treated. The majority of patients (97.5%, n=196) chose to consult MBBS doctors who were registered with the medical board while 2.5% (n=5) selected unregistered healthcare providers. The participants visited health facilities because they felt sick (43.3% (n=87)) and when home remedies stopped working (53.7% (n=108)) and after OTC failure (3% (n=6)). The survey results showed that 63.2% (n=127) participants understood TB becomes fatal when left untreated and 52.7% (n=106) participants knew the length of treatment. The survey results showed that 58.7% (n=118) participants knew about free treatment for TB but 42.8% (n=86) correctly identified the lungs as the primary affected area. The survey results showed that 89.6% (n=180) of participants chose government hospitals for their healthcare needs instead of private hospitals which 10.4% (n=21) participants selected. The first stage of treatment included sputum microscopy for 53.7% (n=108) patients and X-rays for 38.8% (n=78) patients and symptomatic relief for 7.5% (n=15) patients. The diagnosis process needed one visit for 70.6% (n=142) patients but required two visits for 24.4% (n=49) patients and three visits for 3.9% (n=8) patients and four visits for 1% (n=2) patients. The survey results indicated that 97.5% (n=196) participants showed symptoms which matched “Table 2”.

**Table 2:** Distribution of Study Participants by Knowledge Regarding TB and Pre-Diagnosis Healthcare Utilization (n=201)

| Variable | Category | Frequency (%) |
| --- | --- | --- |
| Smoking Causes TB | No | 153 (76.1) |
|  | Yes | 48 (23.9) |
| Knows Symptoms | No | 50 (24.9) |
|  | Yes | 151 (75.1) |
| TB is Infectious | No | 44 (21.9) |
|  | Yes | 157 (78.1) |
| TB is Curable | No | 61 (30.4) |
|  | Yes | 140 (69.4) |
| Treatment is Free | No | 83 (41.3) |
|  | Yes | 118 (58.7) |
| Most Common Part (Lungs) | No | 115 (57.2) |
|  | Yes | 86 (42.8) |
| Life-Threatening if Untreated | No | 74 (36.8) |
|  | Yes | 127 (63.2) |
| Knows Treatment Duration | No | 95 (47.3) |
|  | Yes | 106 (52.7) |
| Health Facility Visits | Every time sick | 87 (43.3) |
|  | Home remedies fail | 108 (53.7) |
|  | OTC fails | 6 (3) |
| Doctor Type | Registered MBBS | 196 (97.5) |
|  | Non-registered | 5 (2.5) |
| Hospital Choice | Government | 180 (89.6) |
|  | Private | 21 (10.4) |
| First Treatment | Symptomatic | 15 (7.5) |
|  | Sputum Microscopy | 108 (53.7) |
|  | X-ray | 78 (38.8) |
| Visits Before Diagnosis | 1 | 142 (70.6) |
|  | 2 | 49 (24.4) |
|  | 3 | 8 (3.9) |
|  | 4 | 2 (1) |
| Symptoms Present | Yes | 196 (97.5) |
|  | No | 5 (2.5) |

The study found that cough was the most frequent symptom reported by patients since it affected 64.7% of participants (n=130) followed by fever which affected 14.9% of participants (n=30). The rare symptom of pneumonia occurred in 0.5% of cases (n=1). The symptoms of chest pain and weight loss and shortness of breath occurred in 4% (n=8) and 1% (n=2) and 1.5% (n=3) of cases respectively. The medical team diagnosed pulmonary TB in 71.6% (n=144) of patients but found extrapulmonary TB in 28.4% (n=57) of cases. The majority of patients obtained their medications from ASHAs (78.1%, n=157) while hospitals (19.4%, n=39) and shops (1.5%, n=3) and other sources (1%, n=2) provided the remaining medications. The perceived diagnostic delays affected 32.3% (n=65) of patients who linked their delays to self-neglect (24.4%, n=49) and doctor uncertainty (4.5%, n=9) and referral processes (13.9%, n=28). The study results show that discrimination against TB patients occurred in only 0.5% of home cases and 2% of community cases and 1% of facility cases “Table 3”.

**Table 3:** Distribution of Study Participants by Symptoms, Diagnosis, Treatment, and Discrimination (n=201)

| Variable | Category | Frequency (%) |
| --- | --- | --- |
| Fever | No | 171 (85.1) |
|  | Yes | 30 (14.9) |
| Cough | No | 71 (35.3) |
|  | Yes | 130 (64.7) |
| Pain and Weakness | No | 192 (95.5) |
|  | Yes | 9 (4.5) |
| Vomiting | No | 184 (91.5) |
|  | Yes | 17 (8.5) |
| Weight Loss | No | 199 (99) |
|  | Yes | 2 (1) |
| Loss of Appetite | No | 195 (97) |
|  | Yes | 6 (3) |
| Haemoptysis | No | 197 (98) |
|  | Yes | 4 (2) |
| Chest Pain | No | 193 (96) |
|  | Yes | 8 (4) |
| Stomach Pain | No | 188 (93.5) |
|  | Yes | 13 (6.5) |
| Enlarged Nodes | No | 184 (91.5) |
|  | Yes | 17 (8.5) |
| Shortness of Breath | No | 198 (98.5) |
|  | Yes | 3 (1.5) |
| Pneumonia | No | 200 (99.5) |
|  | Yes | 1 (0.5) |
| Diagnosis | Pulmonary TB | 144 (71.6) |
|  | Extrapulmonary TB | 57 (28.4) |
| Drug Source | Hospital | 39 (19.4) |
|  | ASHA | 157 (78.1) |
|  | Shop | 3 (1.5) |
|  | Others | 2 (1) |
| Perceived Delay | No | 136 (67.7) |
|  | Yes | 65 (32.3) |
| Delay Reason | Self-neglect | 49 (24.4) |
|  | Doctor uncertainty | 9 (4.5) |
|  | Lack of access | 1 (0.5) |
|  | Increased referrals | 28 (13.9) |
|  | Others | 114 (56.7) |
| Non-Medical Interventions | No | 120 (59.7) |
|  | Yes | 81 (40.3) |
| Sputum Disposal | Bottle and throw | 17 (8.5) |
|  | Flush | 16 (7.9) |
|  | Closed container | 5 (2.5) |
|  | Sputum tray | 3 (1.5) |
|  | Washbasin/running water | 83 (41.2) |
|  | No sputum TB | 62 (30.9) |
|  | Not advised | 15 (7.5) |
| Preventive Measures | Mask | 136 (67.7) |
|  | Isolation | 7 (3.5) |
|  | Both | 4 (2) |
|  | None | 54 (26.8) |
| Treatment Costs | No | 113 (56.2) |
|  | Yes | 88 (43.8) |
| Nikshay Payment | No | 71 (35.3) |
|  | Partial | 76 (37.8) |
|  | Complete | 54 (26.9) |
| Discrimination at Home | No | 200 (99.5) |
|  | Yes | 1 (0.5) |
| Discrimination in Community | No | 197 (98) |
|  | Yes | 4 (2) |
| Discrimination at Facility | No | 199 (99) |
|  | Yes | 2 (1) |

The research reveals multiple vital findings about TB patients’ selection processes for their healthcare providers. People who grasped how smoking leads to TB development and those who understood TB transmission patterns demonstrated better health-seeking practices (OR: 2.33, 95% CI: 1.14-4.74, p=0.01). The patients who used OTC remedies before visiting a doctor for medical help demonstrated superior health-seeking behavior (OR: 13.07, 95% CI: 6.63-25.7, p<0.001). The patients who chose private hospitals over government facilities demonstrated superior health-seeking behavior (OR: 7.94, 95% CI: 1.79-35.13, p<0.001) which suggests that healthcare accessibility and quality perceptions affect their behavior. The research shows that patients choose their healthcare search paths based on the diagnostic procedures and treatment methods they receive. The health-seeking behavior of patients with extrapulmonary TB proved superior to those with pulmonary TB (OR: 2.29, 95% CI: 1.18-4.45, p=0.01). People who recognize the duration between symptom onset and diagnosis receipt want to improve healthcare services (OR: 3.06, 95% CI: 1.58-5.90, p<0.001). The patients who obtain their medication from ASHA workers have lower odds of demonstrating good HSB (OR: 0.33, 95% CI: 0.15-0.75, p=0.01) because they tend to choose traditional healthcare facilities.

The research also reports that patients base their healthcare decisions through a mix of monetary factors and social aspects. The study results show that patients who paid for their TB treatment developed better health-seeking behaviors (OR: 2.75, 95% CI: 1.52-4.97, p<0.001) which indicates that patients who pay for their care become more involved in their health management. The patients who received non-medical intervention advice demonstrated significantly better health-seeking behavior (OR: 5.3, 95% CI: 2.75-10.23, p<0.001) “Table 4”

**Table 4:** Logistic Regression Analysis of Health Seeking behaviour among TB patients with selected variables (n=201)

| Variable | Poor<br>HSB (n) | Good<br>HSB (n) | OR (95% CI) | p-value |
| --- | --- | --- | --- | --- |
| **Age** |  |  |  |  |
| ≤35 years | 49 | 65 | 1 |  |
| >35 years | 35 | 52 | 1.12 (0.63-1.97) | 0.69 |
| **Sex** |  |  |  |  |
| Male | 45 | 62 | 1 |  |
| Female | 39 | 55 | 1.02 (0.58-1.79) | 0.93 |
| **Education Level** |  |  |  |  |
| Illiterate | 11 | 13 | 1 |  |
| Up to 5<sup>th</sup> | 7 | 16 | 1.93 (0.58-6.4) | 0.37 |
| 6<sup>th</sup>-10<sup>th</sup> | 36 | 40 | 0.94 (0.37-2.36) | 0.99 |
| 11<sup>th</sup>-12<sup>th</sup> | 18 | 29 | 1.36 (0.50-3.68) | 0.61 |
| Graduate or Higher | 12 | 19 | 1.33 (0.45-3.94) | 0.78 |
| **Type of Family** |  |  |  |  |
| Nuclear | 53 | 70 | 1 |  |
| Joint | 31 | 47 | 1.14 (0.64-2.0) | 0.63 |
| **Present history of using smokeless tobacco** |  |  |  |  |
| No | 73 | 105 | 1 |  |
| Yes | 11 | 12 | 0.75 (0.31-1.81) | 0.53 |
| **Past history of using smokeless tobacco** |  |  |  |  |
| No | 63 | 82 | 1 |  |
| Yes | 21 | 35 | 1.28 (0.68-2.41) | 0.44 |
| **Present history of using smoke tobacco** |  |  |  |  |
| No | 74 | 103 | 1 |  |
| Yes | 10 | 14 | 1.00 (0.42-2.38) | 0.98 |
| **Past history of using smoke tobacco** |  |  |  |  |
| No | 60 | 77 | 1 |  |
| Yes | 24 | 40 | 1.29 (0.70-2.38) | 0.39 |
| **Present history of using alcohol** |  |  |  |  |
| No | 78 | 106 | 1 |  |
| Yes | 6 | 11 | 1.34 (0.47-3.80) | 0.56 |
| **Past history of using alcohol** |  |  |  |  |
| No | 65 | 88 | 1 |  |
| Yes | 19 | 29 | 1.12 (0.58-2.18) | 0.72 |
| **Knows that smoking causes TB** |  |  |  |  |
| No | 71 | 82 | 1 |  |
| Yes | 13 | 35 | 2.33 (1.14-4.74) | **0.01** |
| **Knows that TB is an infectious disease** |  |  |  |  |
| No | 27 | 17 | 1 |  |
| Yes | 57 | 100 | 2.78 (1.39-5.54) | **<0.001** |
| **Health-Seeking Behavior** |  |  |  |  |
| Every time I feel sick | 64 | 23 | 1 |  |
| Visits Health Facility Only When OTC/Home Remedy Fails | 20 | 94 | 13.07 (6.63-25.7) | **<0.001** |
| **Type of Preferred Healthcare Provider** |  |  |  |  |
| Visits Registered MBBS Doctor | 84 | 112 | - |  |
| Visits Non-Registered MBBS Doctor | 0 | 5 | 0.12 (0.006-2.22) | 0.076 |
| **Choice of Health Facility** |  |  |  |  |
| Prefers Government Hospital | 82 | 98 | 1 |  |
| Prefers Private Hospital | 2 | 19 | 7.94 (1.79-35.13) | **<0.001** |
| **First treatment provided** |  |  |  |  |
| First Advised Symptomatic Treatment | 6 | 9 | 1 |  |
| First Advised Sputum Microscopy/X-Ray | 78 | 108 | 0.92 (0.31-2.69) | 0.88 |
| **Number of visits before diagnosis** |  |  |  |  |
| Number of Visits Before Diagnosis ($\leq 2$) | 84 | 107 | - | |
| Number of Visits Before Diagnosis ($> 2$) | 0 | 10 | 0.06 (0.003-0.97) | **<0.001** |
| **Symptoms before diagnosis** |  |  |  |  |
| Symptoms Present Before Diagnosis | 81 | 115 | 1 |  |
| No Symptoms Before Diagnosis | 3 | 2 | 0.46 (0.07-2.87) | 0.40 |
| **Diagnosis** |  |  |  |  |
| Pulmonary TB | 68 | 76 | 1 |  |
| Extrapulmonary TB | 16 | 41 | 2.29 (1.18-4.45) | **0.01** |
| **Perceived Delay between Symptoms occurrence and diagnosis** |  |  |  |  |
| No | 68 | 68 |  |  |
| Yes | 16 | 49 | 3.06 (1.58-5.90) | **<0.001** |
| **Place of collection of drugs for TB** |  |  |  |  |
| Hospital | 9 | 30 | 1 |  |
| ASHA | 74 | 83 | 0.33 (0.15-0.75) | **0.01** |
| Shop | 1 | 2 | 0.60 (0.22-1.65) | 0.98 |
| Others | 0 | 2 | 0.64 (0.02-14.58) | 0.99 |
| **Were you advised any nonmedical interventions** |  |  |  |  |
| No | 68 | 52 | 1 |  |
| Yes | 16 | 65 | 5.3 (2.75-10.23) | **<0.001** |
| **Have you been using any preventive measures** |  |  |  |  |
| Yes | 67 | 80 | 1 |  |
| No | 17 | 37 | 1.82 (0.94-3.52) | 0.06 |
| **Did you spend any money on treatment of TB** |  |  |  |  |
| No | 59 | 54 | 1 |  |
| Yes | 25 | 63 | 2.75 (1.52-4.97) | **<0.001** |
| **Did you receive any payment from schemes** |  |  |  |  |
| No | 32 | 39 | 1 |  |
| Yes | 52 | 78 | 1.23 (0.68-2.20) | 0.48 |
| **Discrimination at home** |  |  |  |  |
| No | 84 | 116 |  |  |
| Yes | 0 | 1 | 0.46 (0.01-11.42) | 1 |
| **Discrimination at community level** |  |  |  |  |
| No | 83 | 114 |  |  |
| Yes | 1 | 3 | 2.18 (0.22-21.37) | 0.478 |
| **Discrimination at facility level** |  |  |  |  |
| No | 84 | 116 | 1 |  |
| Yes | 0 | 1 | 0.71(0.04-11.6) | 0.814 |

### Qualitative Findings

The study includes qualitative research because health-seeking behavior and TB-related barriers exist as complex systems which need more than quantitative data to understand. The findings show how patients experience TB care through their entire process from detecting symptoms to following treatment instructions and attending scheduled appointments. The stories demonstrate how people make decisions through mental operations and emotional reactions which get influenced by their surroundings and cultural heritage. The structured questionnaires fail to detect the various reasons patients delay their first contact and their family screening methods and their medication routines which appear identical at first glance. The research requires qualitative data to study how patients make decisions and understand their disease and psychological barriers which affect their ability to detect TB symptoms early and finish treatment properly. The qualitative research method provides essential background information to quantitative results which helps developers create patient-focused interventions and improve counseling methods and community-based TB elimination programs “Table 5”.

**Table 5.** Thematic Matrix of Patient Experiences and Responses Throughout the TB Care Cascade.

| Theme | Subtheme | Patient Responses |
| --- | --- | --- |
| Pre-Diagnosis | Discovery of TB | “I went to the hospital because I used to cough a lot in the morning...” |
|  |  | “My fever was not subsiding, so I went to the hospital to get tested...” |
|  |  | “My child’s fever was not reducing, so I went to the hospital...” |
|  | Symptoms Before Testing | Cough, fever, weight loss, fatigue were common pre-diagnosis symptoms. |
| Diagnosis & Treatment | Financial Burden | “I have got all my tests done from Jaipur in a private hospital...” |
|  | Free Treatment Availability | “Everything (tests and medication) was free.” |
|  | Government Support | “ASHA/Health Worker provided all medications free of charge.” |
|  | Medication Timing | “I took the medicines whenever I remembered.” |
|  |  | “Every day at the same time with breakfast.” |
|  |  | “At bedtime.” |
|  |  | "Every day at the same time in the morning." |
|  | Adherence Issues | Some patients reported forgetting doses occasionally. |
|  | Family Screening | "Yes, all family members were screened." |
|  |  | "Only my children were screened." |
|  |  | "No, no one in the family was screened." |
|  | Healthcare Follow-up | "I got a call from the hospital, but they did not visit my home." |
|  | Awareness of Side Effects | "No one told me about the side effects." |
|  |  | "I was informed that my urine colour would change." |
|  |  | "I checked from the internet for information." |
|  | Sputum Disposal Methods | "I throw it in running water." |
|  |  | "Used to flush in the toilet." |
|  |  | "Was not advised about the process of disposal." |

‘Table 6’ shows that in-depth interviews and FGDs reveal crucial information about the concealed barriers which result from stigma and government benefit access and post-treatment healthcare experiences. The quantitative study showed minimal direct discrimination but participants in the qualitative research hid their illness information while staying away from others which blocked their access to medical care and contact tracking. The qualitative research method uncovered three essential issues which quantitative tools failed to detect: Nikshay scheme payment problems and government benefit obstacles and insufficient counseling about side effects and nutrition and sputum disposal. The research results show that quantitative tools cannot detect wide system issues because they stay within predefined limits. The qualitative research method generates essential program management data which shows particular weaknesses in patient education and staff communication and scheme delivery systems. The qualitative research approach confirms quantitative results by creating particular interventions which boost rural TB service delivery.

**Table 6.** Thematic Matrix of Stigma, Access to Government Benefits, and Post-Treatment Healthcare Experiences Among TB Patients.

| Theme | Subtheme | Patient Responses |
| --- | --- | --- |
| Stigma & Discrimination | Family Support | "Everything is fine, no one made me feel discriminated." |
|  | Social Stigma | "I did not inform many family members because they might get scared." |
|  | Community Reactions | "Yes, the neighbourhood feels scared to talk to me." |
| Challenges in Government Benefits | Financial Support Issues | "No, did not receive." |
|  | Partial Benefits | "Received partially." |
|  | Successful Disbursement | "Yes, received till date." |
|  | Lack of Awareness | "Didn't check." |
| Post-Treatment Healthcare Experiences | Recommendations | "Hospitals should provide a pamphlet regarding nutrition in tuberculosis." |
|  | Patient Experience | "Patient is already scared; hospital staff should be polite." |
|  | Doctor-Patient Interaction | "Doctors should give proper time to patients." |
|  | Medicine Availability | "Should provide medicines, and if not available, write alternate medicine." |

## Discussion

The research investigates health-seeking behavior (HSB) among TB patients in rural Gurugram Haryana through a mixed-methods approach which reveals barriers and enablers that affect TB diagnosis and treatment. The study uses statistical data from 201 participants together with qualitative results from five focus group discussions to create vital information which will support the National TB Elimination Program (NTEP) in reaching its 2025 TB elimination target.

The study shows that 58.2% of participants showed good health-seeking behavior because they understood TB is contagious (OR 2.78, p=0.003) and smoking leads to TB development (OR 2.33, p=0.015). The research supports Hussain et al [5] who demonstrated that tribal Odisha TB patients achieved better results through their education and health awareness. The research results showed that participants had limited knowledge about smoking risks for TB development at 23.9% yet they understood TB symptoms and infectiousness at 75.1% and 78.1% respectively. The research indicates that health education reaches different levels of the population because people understand symptoms better than they understand smoking as a TB risk factor. The qualitative results indicate that patients tend to view their cough as non-threatening until their symptoms advance to include fever and weight loss.

The research demonstrates that health campaigns need to focus on teaching people about risk factors and symptoms because rural areas show high smoking rates of 11.9% current and 31.9% past [7]The study shows that most participants (89.6%) chose to receive their care at government hospitals and 97.5% of them selected registered MBBS doctors for treatment. The study shows that patients who chose private facilities for their care demonstrated better health-seeking behavior (OR 7.94, p=0.0006). The patients chose private facilities because they believed these facilities provided better quality care and shorter treatment times. The qualitative data indicates that Jaipur patients chose to spend money on private medical tests because they wanted to receive treatment in urban areas near Gurugram despite NTEP offering free services.

Diagnostic delays, reported by 32.3%, with self-neglect (24.4%) and referrals (13.9%) as primary reasons, mirror findings by Sreerama Reddy et al [8], who identified patient and system delays as key hurdles in India. The quantitative association with poor HSB (OR 3.06, p=0.0005) and qualitative accounts of symptom dismissal (“thought it was normal”) indicate a dual challenge: patient hesitancy and healthcare system inefficiencies. Notably, 70.6% received a diagnosis in one visit, suggesting that once engaged, the system is efficient for many. However, the 24.4% requiring two visits and 4.9% needing three or more point to bottlenecks—possibly diagnostic capacity or referral pathways—that warrant investigation. Strengthening community-level screening, as 53.7% sought care only after home remedies failed (OR 13.07, p<0.001), could bridge this gap [9]

Despite NTEP’s free treatment promise, 43.8% incurred costs, a significant barrier to good HSB (OR 2.75, p = 0.0006). This echoes findings from Mumbai reporting substantial out-of-pocket expenditures, particularly for diagnostics [10–11]. Qualitative data reveal that costs are concentrated in private tests rather than medications, since 78.1% of patients received drugs via ASHAs [12]. This frontline worker model is a clear enabler—reducing drug procurement barriers (OR 0.33, p = 0.01 for ASHA vs. hospital) yet incomplete Nikshay Poshan Yojana disbursements (37.8% partial, 26.9% complete) suggest implementation gaps [13]. Patients’ calls for nutritional support (“hospital should provide a pamphlet”) highlight unmet needs exacerbated by financial strain, especially among the 42.3% unemployed. Expanding ASHA roles to include nutritional counselling and ensuring full scheme disbursements could mitigate these economic hurdles [14].

Extrapulmonary TB (28.4%) was a barrier to good HSB (OR 2.29, p=0.011), likely due to diagnostic complexity compared to pulmonary TB (71.6%), which typically presents with cough (64.7%). This aligns with global evidence that extrapulmonary cases face longer delays due to atypical symptoms (e.g., only 1% weight loss here) and limited diagnostic tools in rural settings. Cough’s dominance contrasts with lower fever prevalence (14.9%), suggesting symptom-specific education could prioritize respiratory signs [15].

Substance use (e.g., 11.4% smokeless tobacco, 8.5% alcohol) showed no significant HSB impact (p>0.05), differing from studies linking alcohol to TB progression [16]. This may reflect underreporting or a sample-specific low prevalence, necessitating larger studies to clarify behavioral influences. Discrimination was minimal (0.5–2%), contrasting with Deo et al. (2020)’s urban findings of overt stigma. Yet, qualitative data reveal subtle social barriers: fear of disclosure (“didn’t tell my family”) and community avoidance (“neighbours feel scared”). This internalized stigma, less quantifiable but impactful, may delay care-seeking and contact tracing, as only 40.3% received non-medical advice like isolation. Preventive measures (67.7% masks) indicate some awareness, but inconsistent sputum disposal (41.2% washbasins) suggests gaps in infection control education, critical for TB’s airborne transmission Qualitative critiques of staff “doctors are too busy” and calls for politeness resonate with patient-centred care principles [17]. The 59.7% not advised on non-medical interventions and 47.3% unaware of treatment duration reflect counseling deficiencies, potentially undermining adherence. This contrasts with ASHA’s drug delivery success, highlighting a disparity between logistical and educational support. Enhancing provider training and patient engagement tools (e.g., pamphlets) could address these shortcomings, aligning with NTEP’s community mobilization goals. Compared to urban studies^4^, rural Gurugram shows stronger public system engagement but persistent economic and awareness barriers. Post-COVID-19, these align with global trends of disrupted TB care, emphasizing the urgency of adaptive strategies. This study’s mixed-methods approach offers a model for other rural Indian settings, balancing statistical rigor with lived experiences.

These findings inform NTEP’s 2025 elimination strategy. Knowledge gaps require multimedia campaigns on smoking and symptoms, leveraging ASHAs for outreach. Diagnostic delays and costs call for decentralized sputum testing and full Nikshay coverage. Stigma reduction needs community-level sensitization, while healthcare interactions demand staff training in empathy and communication. Rural Gurugram’s proximity to urban facilities offers a unique opportunity to integrate public-private partnerships, reducing private care reliance Recall bias may affect symptom and delay reporting, given the retrospective design. The exclusion of uncontactable patients (e.g., via wrong numbers) risks underrepresenting severe cases or defaulters, potentially skewing HSB scores. The cross-sectional nature limits causality inference, and the small qualitative sample (five FGDs) may miss diverse perspectives. Future longitudinal studies with broader sampling could validate these findings.

## Conclusion

The research conducted in rural Gurugram Haryana through mixed-methods approaches shows how knowledge and healthcare availability and social economic conditions influence TB patients’ health-seeking behavior (HSB) which offers vital information for India’s National TB Elimination Program (NTEP). The study shows that 58.2% of participants achieved excellent HSB results through their TB transmission education and their decision to visit doctors after home treatments did not work (OR 2.78, p=0.003) (OR 13.07, p<0.001). The research demonstrates that educational programs and outreach activities lead to better early disease detection and improved patient compliance with treatment. The study shows that multiple obstacles continue to exist because patients experience shows patients delay their diagnosis by 32.3% due to their own self-care failures and healthcare system problems and they must pay 43.8% of their medical expenses out of pocket even though NTEP provides free services which mainly affect the 42.3% unemployed population. The research results show that patients face hidden discrimination because healthcare staff do not give enough guidance and patients do not follow preventive measures because they cannot properly dispose of their sputum (41.2%). The study shows that ASHAs deliver 78.1% of drugs and patients choose government facilities at 89.6% but these advantages suffer from poor understanding of risk elements including smoking (23.9%) and insufficient Nikshay scheme funding (only 26.9% receive full payments). The research shows that rural Gurugram can use NTEP infrastructure but TB elimination in 2025 requires complete solutions which need to address multiple challenges.

## Recommendations

The implementation of a comprehensive strategy will help HSB to become stronger while achieving TB elimination goals. The NTEP needs to create specific multimedia programs which teach TB risk elements and warning signs through ASHAs and community health workers to reach the 76.1% of people who do not understand smoking causes TB and to enhance the 75.1% who recognize symptoms. The implementation of sputum microscopy and X-ray services at village health posts will decrease diagnostic delays to 32.3% while supporting the 70.6% of patients who receive their diagnosis during their first visit. The training program for ASHAs needs to include instruction about patient side effects and infection control measures including sputum disposal methods that go beyond the current 41.2% who use washbasins. The Nikshay Poshan Yojna needs complete execution to achieve 100% coverage which exceeds the present 26.9% level while adding nutritional support according to FGD recommendations to reduce the 43.8% of treatment expenses. Healthcare providers who receive training about patient-centered communication methods will learn to handle patient complaints about staff conduct which will establish trust and enhance treatment compliance. The research recommendations from quantitative and qualitative data support NSP 2017–25 targets by creating a method to eliminate TB from rural India through enhanced system efficiency and removal of critical barriers.

## Data Availability

The data is available with the researchers and can be shared through a proper channel in the event of a requirement or if the paper is accepted.

## Acknowledgement

The research team wish to show the gratitude to the TB patients who participated the study and provided their valuable information and recommendations for the strengthening of the program

## References

1. World Health Organization. Global Tuberculosis Report 2021. Geneva: WHO; 2021.

2. Ministry of Health and Family Welfare. National Strategic Plan for TB Elimination 2017–2025. New Delhi: Government of India; 2017.

3. Deo S, Jha N, Mohanty S, Sabde Y, Dharmshaktu NS. Barriers to tuberculosis diagnosis and treatment in India: A systematic review. Indian J Tuberc. 2020;67(1):12–21.

4. Mistry N, Rangan S, Dholakia Y, Lobo E, Shah S, Patil A, et al. Challenges and opportunities for engaging the private sector in tuberculosis care in India. PLoS One. 2017;12(9):e0184686.

5. Hussain T, Sinha A, Nagaraja SB, Tullu FT, Parmar MC, Mehta KG, et al. Impact of health education on tuberculosis awareness and treatment outcomes in tribal Odisha, India. Indian J Community Med. 2020;45(3):286–92.

6. Slama K, Chiang CY, Enarson DA, Hassmiller K, Fanning A, Gupta P, et al. Tobacco and tuberculosis: A qualitative systematic review and meta-analysis. Int J Tuberc Lung Dis. 2007;11(10):1049–61.

7. Global Adult Tobacco Survey Collaborating Group. Global Adult Tobacco Survey (GATS) India 2016–17. Mumbai: Tata Institute of Social Sciences; 2018.

8. Sreeramareddy CT, Qin ZZ, Satyanarayana S, Subbaraman R, Pai M. Delays in diagnosis and treatment of pulmonary tuberculosis in India: a systematic review. Int J Tuberc Lung Dis. 2014;18(3):255–66. doi:10.5588/ijtld.13.0585

9. World Health Organization. WHO operational handbook on tuberculosis: module 2: screening – systematic screening for tuberculosis disease. Geneva: World Health Organization; 2022.

10. Satyanarayana S, Kumar S, Bhavsar A, et al. Engaging the private health service delivery sector for tuberculosis care in India: a cost analysis. Trop Med Infect Dis. 2023;8(5):265.

11. Shetty N, Rodrigues R, Dhar A, et al. Health-seeking behaviour of drug-resistant tuberculosis patients in Mumbai, India: a qualitative study. PLoS One. 2018;13(3):e0194428.

12. Lokuge KM, Verma G, John KR, et al. Role of community health workers in improving cost efficiency in tuberculosis treatment delivery: evidence from India. BMC Public Health. 2020;20:123.

13. Nikshay Poshan Yojana. Indian government nutritional support scheme for TB patients. Wikipedia. Updated February 2025.

14. National Health Systems Resource Centre. ASHA Update Jan 2018. New Delhi: NHSRC; 2018.

15. Purohit MR, Mustafa T. Extrapulmonary tuberculosis: pathogenesis and diagnostic approaches. Int J Mycobacteriol. 2015;4(4):221–227.

16. Imtiaz S, Shield KD, Roerecke M, Samokhvalov AV, Rehm J. Alcohol consumption as a risk factor for tuberculosis: meta-analyses and burden of disease. Eur Respir J. 2017;50(1):1700216.

17. Epstein RM, Street RL Jr. The values and value of patient-centered care. Ann Fam Med. 2011;9(2):100–103.

